# A customizable multiplex protein microarray for antibody testing and its application for tick-borne and other infectious diseases

**DOI:** 10.1101/2023.10.06.23296637

**Authors:** Hari Krishnan Krishnamurthy, Vasanth Jayaraman, Karthik Krishna, Tianhao Wang, Kang Bei, Chithra Suresh, Shiny Matilda, Alex J Rai, Renata Welc-Falęciak, Agnieszka Pawełczyk, Lucas S. Blanton, Aleš Chrdle, Andrea Fořtová, Daniel Růžek, Gheyath K. Nasrallah, Laith J. Abu-Raddadi, Duaa W. Al-Sadeq, Marah Abed Alhakim Abdallah, Daniele Lilleri, Chiara Fornara, Piera D’Angelo, Milena Furione, Maria Söderlund- Venermo, Klaus Hedman, Dimosthenis Chochlakis, Anna Psaroulaki, Eirini Makridaki, Artemis Ntoula, John J. Rajasekaran

**Affiliations:** Vibrant Sciences LLC., San Carlos, CA, United States of America; Vibrant America LLC., San Carlos, CA, United States of America; Columbia University, Irving Medical Center, Dept. of Pathology & Cell Biology; Department of Parasitology, Faculty of Biology, University of Warsaw, Diagnostic Laboratory of Parasitic Diseases and Zoonotic Infections, Biological and Chemical Research Centre, Warsaw, Poland; Department of Immunopathology of Infectious and Parasitic Diseases, Medical University of Warsaw, 3C Pawińskiego Street, 02-106, Warsaw, Poland; Department of Internal Medicine, Division of Infectious Diseases, University of Texas Medical Branch, Galveston, TX 77555, USA; Department of Infectious Diseases, Hospital Ceske Budejovice, Ceske Budejovice, Czech Republic; Royal Liverpool University Hospital, Prescot St, Liverpool L7 8XP, UK; Veterinary Research Institute, Brno, Czech Republic; Faculty of Science, Masaryk University, Brno, Czech Republic; Institute of Parasitology, Biology Centre of the Czech Academy of Sciences, Ceske Budejovice, Czech Republic; Veterinary Research Institute, Brno, Czech Republic, Faculty of Science, Masaryk University, Brno, Czech Republic; Biomedical Sciences Department, College of Health Sciences, Qatar University, Doha P.O. Box 2713, Qatar; Microbiologia e Virologia, Fondazione IRCCS Policlinico San Matteo, 27100 Pavia, Italy; Virology, University of Helsinki, Haartmaninkatu 3, FI-00290, Helsinki, Finland; Laboratory of Clinical Microbiology and Microbial Pathogenesis, School of Medicine, University of Crete, PC 70013, Heraklion – Crete – Greece

**Keywords:** Tick-borne infections, Lyme disease, multiplex, microarray, immunoglobulin, tick bite, co-infections, infectious disease

## Abstract

Tick-borne infections are the most common vector-borne diseases in the USA. Ticks harbor and spread several infections with Lyme disease being the most common tickborne infection in the US and Europe. Lack of awareness about tick populations, specific diagnostic tests, and overlapping symptoms of tick-borne infections can often lead to misdiagnosis affecting treatment and the prevalence data reported especially for non-Lyme tick-borne infections. The diagnostic tests currently available for tick-borne diseases are severely limited in their ability to provide accurate results and cannot detect multiple pathogens in a single run. The multiplex protein microarray developed at Vibrant was designed to detect multiple serological antibodies thereby detecting exposure to multiple pathogens simultaneously. Our microarray in its present form can accommodate 400 antigens and can multiplex across antigen types, whole cell sonicates, recombinant proteins, and peptides. A designed array containing multiple antigens of several microbes including *Borrelia burgdorferi,* the Lyme disease spirochete, was manufactured and evaluated. The immunoglobulin M (IgM) and G (IgG) responses against several tick-borne microbes and other infectious agents were analyzed for analytical and clinical performance. The microarray improved IgM and IgG sensitivities and specificities of individual microbes when compared with the respective gold standards. The testing was also performed in a single run in comparison to multiple runs needed for comparable testing standards. In summary, our study presents a flexible multiplex microarray platform that can provide quick results with high sensitivity and specificity for evaluating exposure to varied infectious agents especially tick-borne infections.

## Introduction

Most vector-borne infections in the USA can be attributed to pathogens transmitted via tick bites. Of all tick-borne infections identified to date, Lyme disease is the most prevalent infection [1]. Lyme disease is a potentially serious bacterial infection transmitted by ticks and was first reported in the mid-1970s in the USA. The etiological agent was identified later as *Borrelia burgdorferi* [2,3,4]. Several studies have reported the presence of co-infections along with Lyme disease [5] including *Babesia spp.* [6]*, Bartonella spp.* [7]*, Ehrlichia spp.* [8]*, Anaplasma phagocytophilum* [8], Powassan Virus [9], *Toxoplasma gondii* [10]*, Rickettsia spp.* [11], tick-borne encephalitis virus [12], and West Nile virus [13]. Additionally, prolonged exposure to Lyme and other tick-borne infections could potentially weaken the patient’s immune system increasing the risk of infections like Epstein Barr virus [14], cytomegalovirus [14], parvovirus B19 [5], coxsackie virus [15], HSV-1 [16], HSV-2 [16], and HHV-6 [14].

Ticks have been shown to transmit more than one infectious agent in a single bite. For instance, a study by Wormser et al. showed that there was a chance of getting infected with *A. phagocytophilum* (30%) and *B. microti* (24%) along with Lyme disease [17]. Currently, multi-tiered testing is carried out for diagnosing tick-borne infections [18] (https://tinyurl.com/yeyxevve). In this method, the infectious agents are tested sequentially, starting with Lyme disease. This method is time-consuming and can often lead to delayed diagnosis, accompanied with high cost to the patient [18, 19] (https://tinyurl.com/yeyxevve). Testing for multiple infections in a single run can help physicians arrive at an accurate diagnosis especially since Lyme disease shares symptoms with other vector-borne co-infections [20]. The existing diagnostic assays possess various limitations that restrict their applicability in the diagnosis of these infections. The diagnosis of Lyme disease and other infections using several blot-based and single-plex ELISA tests remain rudimentary in terms of arriving at a diagnostic conclusion [21]. Additionally, blot-based assays may have overlapping proteins with similar mass requiring additional testing to tease out the specific antigen to which the antibody is bound. A multiplex system can detect the biomarkers of Lyme disease, potential co-infections, and other infections in a single run. A serology-based multiplexing system may be preferred to a PCR multiplex system mainly due to its accessibility, for instance using dried blood spots [22]. Additionally, serology overcomes the issue of low availability of genetic material due to the transient nature of some of these organisms [23]. A serological-based system is also ideal for population screening and surveillance since it can indicate past exposure to a pathogen.

Our customisable protein microarray design includes antigens physically separated by design unlike blot assays and can multiplex across species. Multiplexing can also be done across antigen types such as recombinant proteins, peptides, and lysates simultaneously. This method can lower test costs since all the manufacturing is automated using bio customised semiconductor processes similar to how electronic chips are made. The multiplex microarray has three main advantages over the existing technologies. It has an ultra-high-density array surface with high reproducibility and better throughput. It can detect a large number of antibodies against varied infectious agents at the same time. Detection of antibodies can be performed using low sample volumes with low cost and a fast turnaround time [21]. Given the flexible nature of the multiplex platform, we aimed to provide a multiplexed testing solution for Lyme, its co-infections, and other possible infections of interest.

## Materials and Methods

### Patients Sera

The sera from 2990 individuals were collected after seeking appropriate Institutional Review Board (IRB) approval under respective collaborators (Supplementary Table 6). Table 1 lists the provided samples for Lyme disease, co-infections, and other infections along with the counts, respective collaborators and methods used to ascertain the clinical diagnosis by the physician. These reference sera were tested at Vibrant America Clinical Labs (CLIA and CAP accredited facility) by laboratory personnel in a blinded manner. The sera from healthy patients were considered negative and were used to set the cut-off values and were investigated under IRB exemption (work order #1-1574995-1) determined by the Western Institutional Review Board (WIRB) to employ de-linked and de-identified human specimens and medical data for research findings. The negative sera were collected from across the US including endemic and nonendemic regions for these infections.

**Table 1.** Sample Cohort. provides an overview of the pathogens used in the study along with the total number of samples, basis of diagnosis, and sample source.

| Pathogen | N | Source (scientist/company) | Basis of Diagnosis |
| --- | --- | --- | --- |
| *Borrelia burgdorferi* | 298 | CDC, Private Clinics | RT-PCR, Physician |
| *Babesia microti* | 70 | Parasitology Laboratory, Wadsworth Center (NYSDOH) | RT-PCR, Blood Smear |
| *Babesia microti* | 118 | Seracare, Boca Bio, Private Clinics | Serology, RT-PCR |
| *Babesia microti* | 26 | Renata Welc-Falęciak (University of Warsaw), Agnieszka Pawełczyk (Warsaw Medical University) | Physician |
| *Bartonella henselae* | 119 | Private clinics | RT-PCR, Physician |
| *Bartonella henselae* | 26 | Renata Welc-Falęciak (University of Warsaw), Agnieszka Pawełczyk (Warsaw Medical University) | Physician |
| *Bartonella henselae* | 10 | Dimosthenis Chochlakis (University of Crete) | Physician |
| *Anaplasma phagocytophilum* | 118 | Private clinics, Boca Bio | RT-PCR, Serology |
| *Anaplasma phagocytophilum* | 26 | Renata Welc-Falęciak (University of Warsaw), Agnieszka Pawełczyk (Warsaw Medical University) | Physician |
| *Ehrlichia chaffeensis* | 120 | Private clinics, Boca Bio | RT-PCR, Serology |
| *Ehrlichia chaffeensis* | 26 | Renata Welc-Falęciak (University of Warsaw), Agnieszka Pawełczyk (Warsaw Medical University) | Physician |
| *Rickettsia typhi* | 70 | Lucas Blanton ( University of Texas Medical Branch) | Physician |
| *Rickettsia typhi* | 124 | Private clinics | RT-PCR, Physician |
| Powassan virus | 127 | Private clinics | RT-PCR, Physician |
| Tick-borne encephalitis | 111 | Daniel Ružek (Czech Academy of Sciences) | Physician |
| **Tick-borne encephalitis** | 124 | Private clinics | RT-PCR, Physician |
| **West Nile virus** | 20 | Gheyath K. Nasrallah (Weill Cornell Medicine-Qatar) | Physician |
| **West Nile virus** | 124 | Private clinics | RT-PCR, Physician |
| **Coxsackie virus** | 45 | iSpecimen | Serology |
| **Coxsackie virus** | 124 | Private clinics | RT-PCR, Physician |
| **Cytomegalovirus** | 43 | DLS | Serology |
| **Cytomegalovirus** | 138 | Daniele Lilleri (Fondazione IRCCS Policlinico San Matteo) | Physician |
| **Cytomegalovirus** | 96 | Private Clinics | RT-PCR, Physician |
| **Cytomegalovirus** | 37 | Seracare | Serology |
| **Epstein Barr virus** | 20 | Gheyath K. Nasrallah (Weill Cornell Medicine-Qatar) | Physician |
| **Epstein Barr virus** | 43 | Seracare | Serology |
| **Epstein Barr virus** | 14 | iSpecimen | Serology |
| **Epstein Barr virus** | 96 | Private clinics | RT-PCR, Physician |
| **Parvovirus B19** | 124 | Private clinics | RT-PCR, Physician |
| **Toxoplasma gondii** | 24 | Seracare | Serology |
| **Toxoplasma gondii** | 124 | Private clinics | RT-PCR, Physician |
| **HSV-1** | 20 | Gheyath K. Nasrallah (Weill Cornell Medicine-Qatar) | Physician |
| **HSV-1** | 31 | Seracare | Serology |
| **HSV-1** | 96 | Private clinics | RT-PCR, Physician |
| **HSV-2** | 19 | Gheyath K. Nasrallah (Weill Cornell Medicine-Qatar) | Physician |
| **HSV-2** | 27 | Seracare | Serology |
| **HSV-2** | 96 | Private clinics | RT-PCR, Physician |
| **HHV-6** | 20 | Gheyath K. Nasrallah (Weill Cornell Medicine-Qatar) | Physician |
| **HHV-6** | 96 | Private clinics | RT-PCR, Physician |

## Processing of Wafers

Wafers were functionalized as described previously [21,24] (https://tinyurl.com/mr9ctppy). Briefly, silicon wafers were exposed to an environment of pure oxygen for 2h followed by washing (deionized Water) and coating (1% (vol/vol) with 3-aminopropyltriethoxysilane (APTES) in N-methylpyrrolidone (NMP). Curing was carried out at 120 °C for 60 minutes under an N2 atmosphere and humidity-controlled environment. Coating and incubation of the wafer with a co-polymer solution of poly (L-lysine) and poly (lactic acid) for 24h were carried out to increase the binding efficiency of the surface on to which the antigens were immobilized via passive adsorption/hydrophobic interactions with the copolymers [Figure 1].

**Figure 1:**
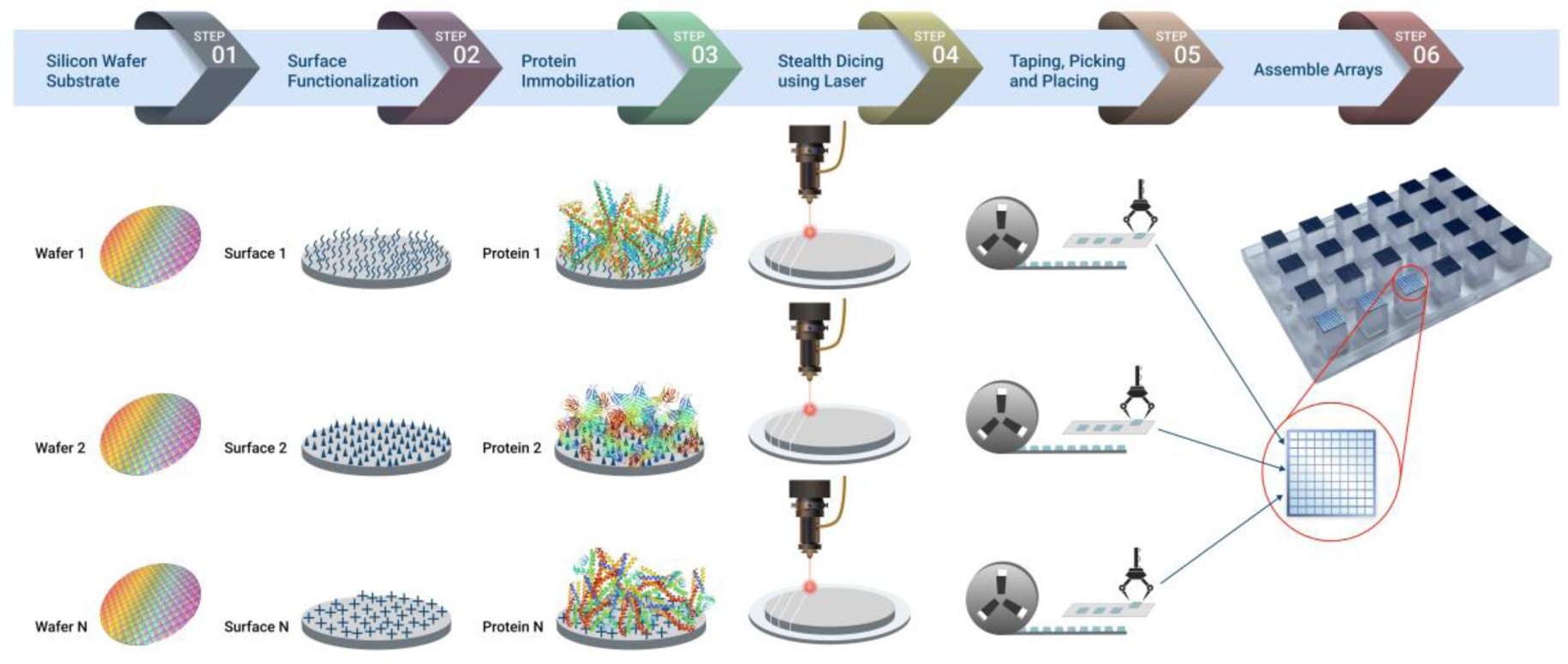
Wafer Processing, Antigen Immobilisation, Pillar Plate Assembly. A poly (lactic acid) and poly (L-lysine) copolymer solution is coated onto the silicon wafers and further immobilized with protein probes [steps 1-3]. The wafers are then diced into microchips using a stealth dicing process [step 4]. A standard die sorting system is used to pick and place the microchips onto carrier plates [step 5]. The carrier tapes are loaded onto a high throughput surface mount technology (SMT) component placement system and individual microchips are placed onto 24-pillar plates. Each pillar consists of 87 microchips [step 6].

## Immobilization of Antigens

The antigens included in the assay are listed in Table 2. Pathogens transmitted by ticks and their respective antigens for potential future additions are listed in Figure 2 [25–30] (https://tinyurl.com/37dprsy7). The recombinant antigens were expressed in *E. coli* bacteria using full-length cDNA coding for the respective antigens fused with a hexa histidine purification tag. The whole cell sonicate was obtained from organisms cultured according to ATCC protocols prior to lysing them which yielded a cocktail of the cell membrane, cell wall, and cytosolic proteins. Peptide antigens were synthesized by photolithography as shown in our previous publications [31,32]. The capture antigens including the recombinant antigens that mimic the natural pathogen and the whole-cell sonicate were incubated on the wafer at a concentration of 1.0 μg/ml and reacted for 24h at 4 °C. The unbound antigens were removed by washing with aqueous phosphate buffer and the unreacted substrate was quenched with a blocking solution containing BSA and glycine. The immobilized antigens were classified with unique identifiers assigned to each wafer. In this study, we employed the microarray to detect Lyme disease, co-infections, and other agents of interest including, *B. microti, B. henselae, A. phagocytophilum, E. chaffeensis, R. typhi,* Powassan virus, tick-borne encephalitis virus, West Nile virus, coxsackie virus, cytomegalovirus, Epstein Barr virus, parvovirus B19, *T. gondii,* HSV-1, HSV-2, and HHV-6 [Figure 2].

**Figure 2:**
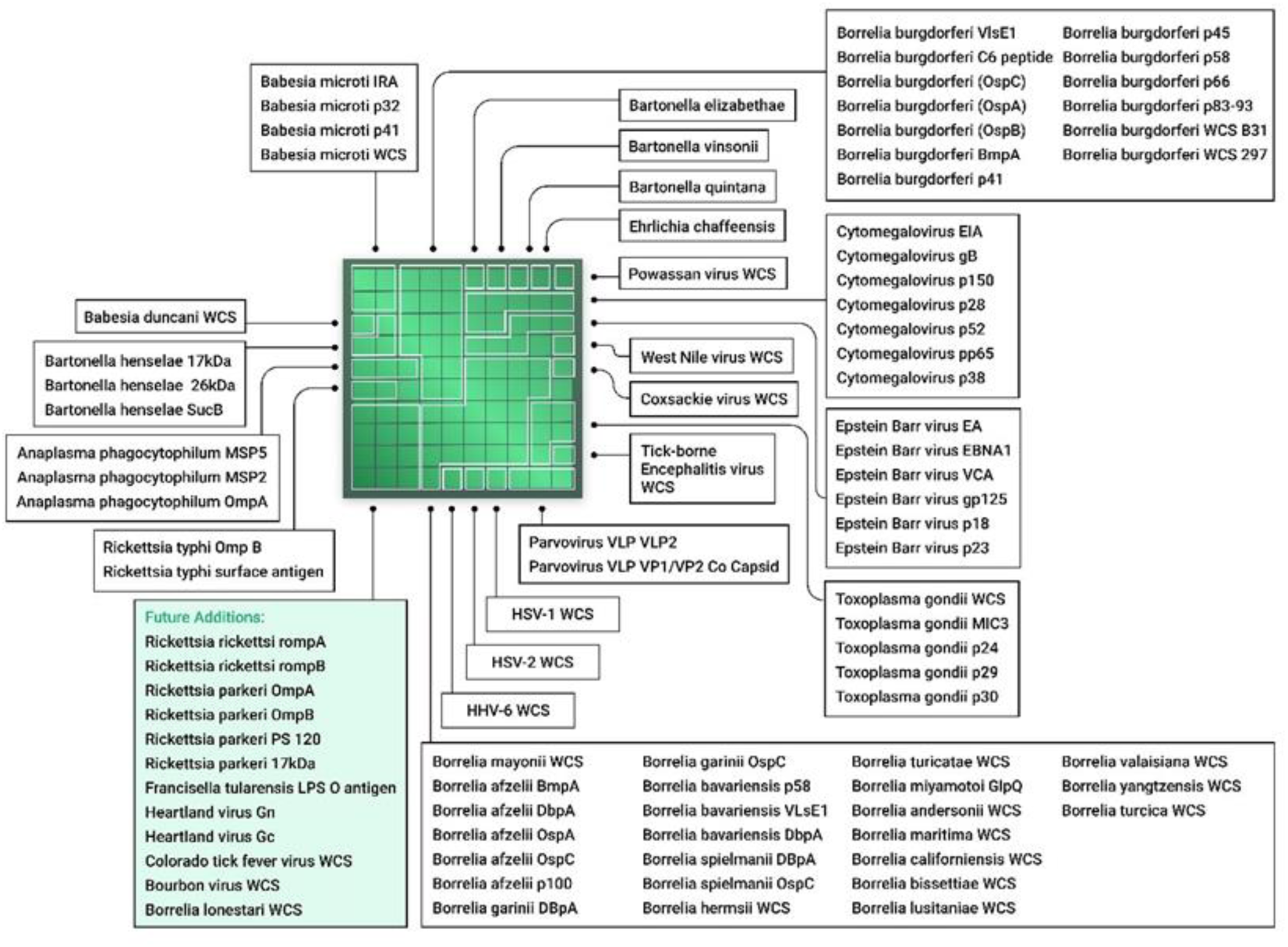
O**v**erview **of all the pathogens and their respective antigens used in the study**.

**Table 2.**
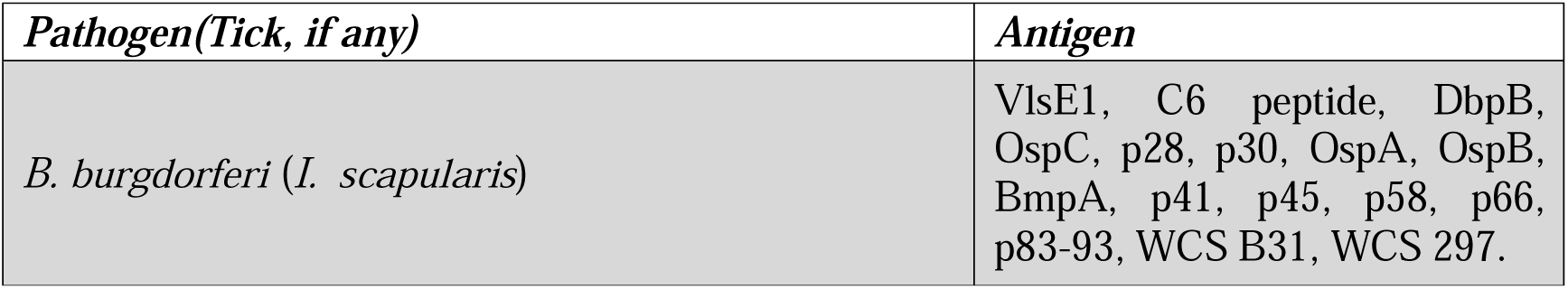
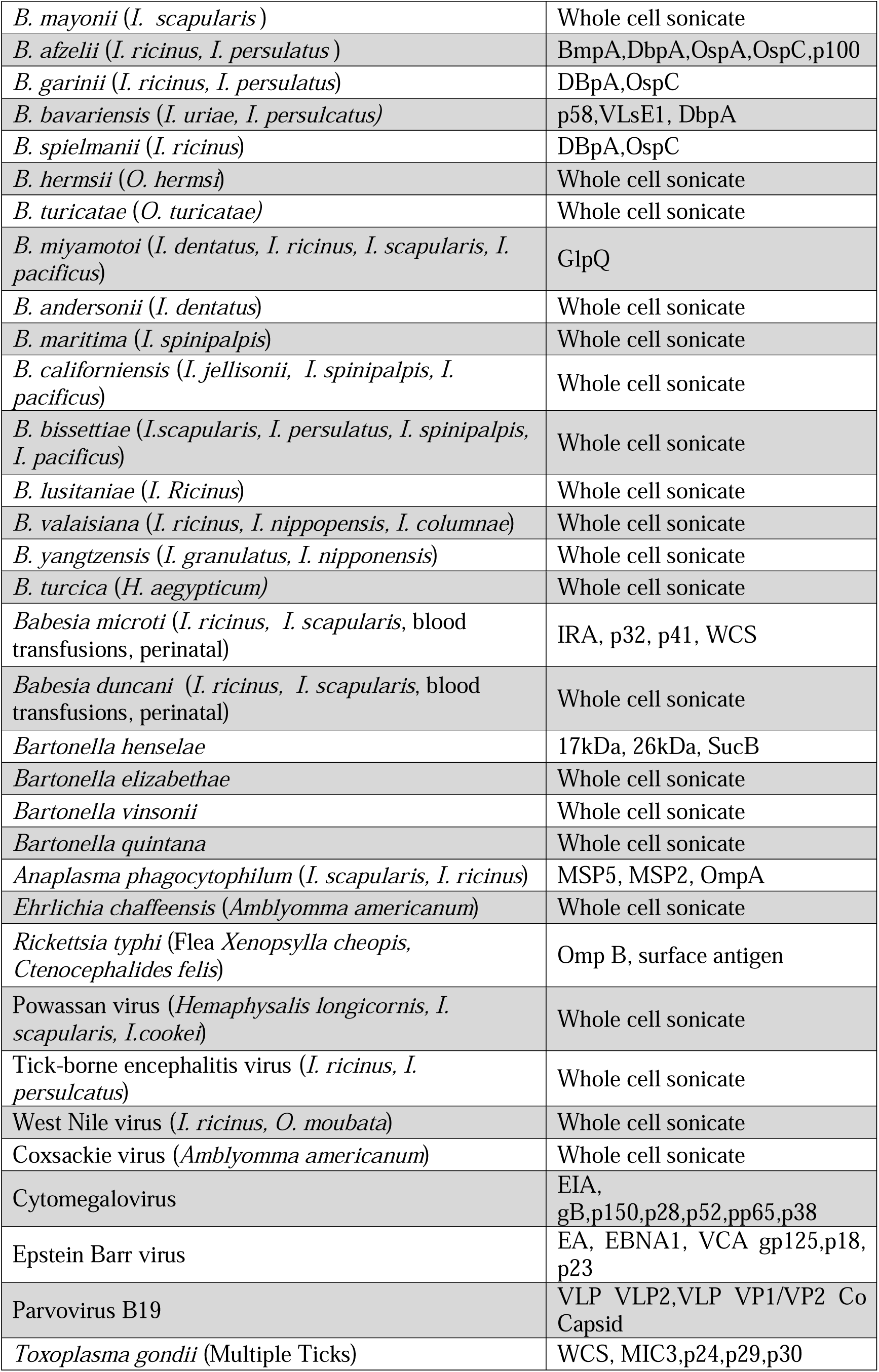
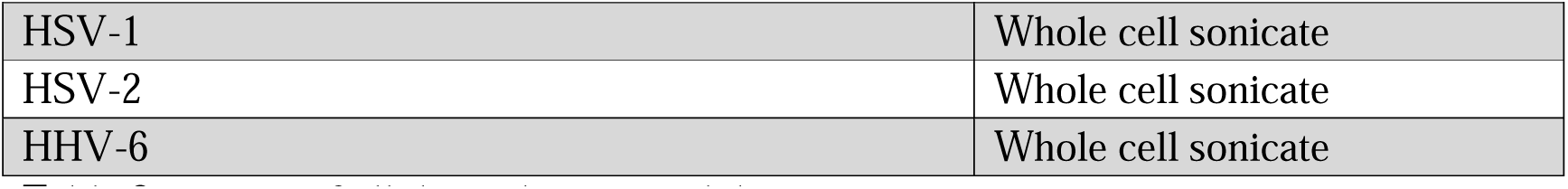
Overview of pathogens and antigens. consists of all the pathogens and their respective antigens.

## Pillar Plate Assembly

Individual wafers were stealth diced into 0.70 x 0.70mm^2^ microchips for each antigen. A standard die-sorting system was used to pick and place these wafers onto individual carrier tapes. The carrier tapes were then placed onto a high-throughput surface mount technology (SMT) component placement system. Finally, microchips were mounted onto 24 pillar plates and each pillar contains 87 microchips with each chip designated for one antigen – recombinant protein, peptide or whole cell sonicates [Figure 1].

## Immunochip assay and Antibody detection

Serum samples were probed using 1:20 dilution on the pillar plate and incubated for 1h at room temperature followed by alternate washing and incubation as described previously [21]. The plate was then incubated for an hour with the secondary antibody (1:2000 dilution of Goat Anti-Human IgG HRP and Goat Anti-Human IgM HRP individually) and washed with TBST buffer followed by DI Water. The plates were left for drying preceding the addition of chemiluminescent substrate and the performance of chemiluminescent imaging. An enhanced IgM sensitivity was achieved by pre-reacting the sera with proprietary assay components leading to IgG stripping prior to IgM testing.

The detection of multiplex antibodies is based on the chemiluminescent immunoassay and can be performed using <200 μL of serum. Sample dilution, multi-step incubation, and multi-solution washing are programmed into liquid handlers. The immunochip has the capacity to assay 192 individual specimens in 2h. Raw chemiluminescent signals for each probe are extracted and converted into intensity plots by an in-house reporter software. This method of automatic antigen detection can dramatically shorten the turnaround time, reduce the cost of labor and instrument, and eliminate the need for manual handling and subjective interpretation of the WB or IB test results when compared to the traditional two-tiered testing recommended by the CDC. All the antibodies are detected in a single run.

## Data analysis

An in-house software extracts the chemiluminescent signals from the generated images which were converted to intensity plots. The average intensity of each antibody was compared with the cut-off values assigned for each antigen to track seropositivity.

## Results

### Custom Protein Microarray Platform

The main components of the Immunochip platform include multiple silicon-based 0.70 ×

0.70 mm^2^ microchips that are laser diced from antigen-immobilized wafers, a customized 24 well compatible plate containing 24 pillars, each containing 87 microchips that are picked and placed into a multiplex microarray assembly, and a high-resolution imager capable of simultaneously detecting chemiluminescent signals from labelled antigen–antibody reactions at each microchip throughout the multiplex microarray (Figure 1). Each chip can be considered analogous to an individual band in a Western blot; however, the proteins are physically separated eliminating cross-reactive issues usually seen in blot-based assays for proteins with similar mass. Figure 1 provides an overview of the microarray manufacturing process. Figure 2 shows the individual chips that are placed in each pillar, a single serum sample will be applied to each pillar thereby assaying the antibodies in serum against all antigens at the same time.

## Analysis of serological response

The Vibrant tick-borne disease panel tests for IgG and IgM antibodies for Lyme disease and other infectious agents as mentioned in Table 2 and Figure 2. The IgM and IgG immune responses were analysed, and the clinical sensitivities and specificities were tabulated in Table 3. The samples reacted with a specific immunoreactive epitope of the 87 different antigens that were being tested. The immunoreactivity of these antigens was contrasted with that of the controls.

**Table 3.**
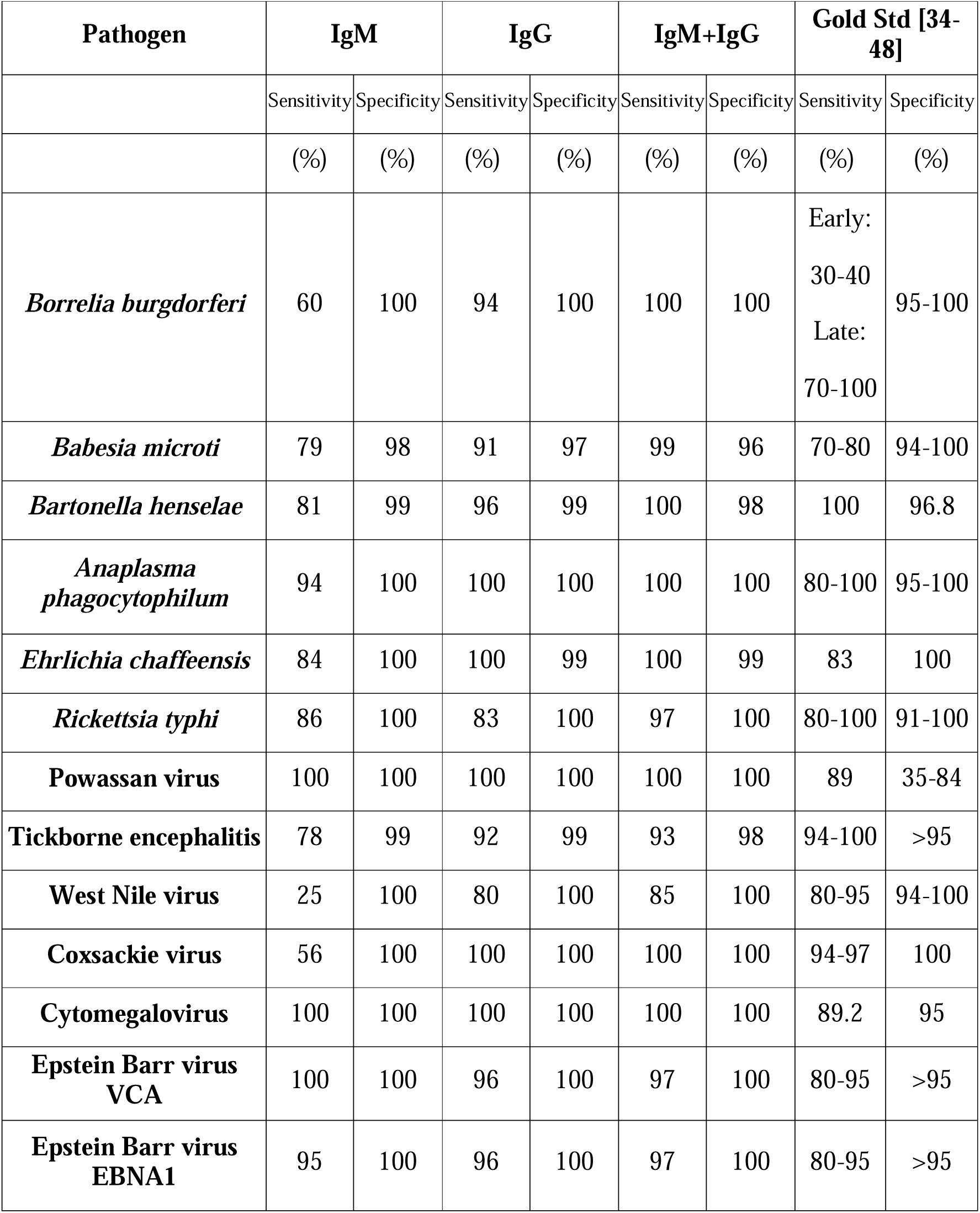
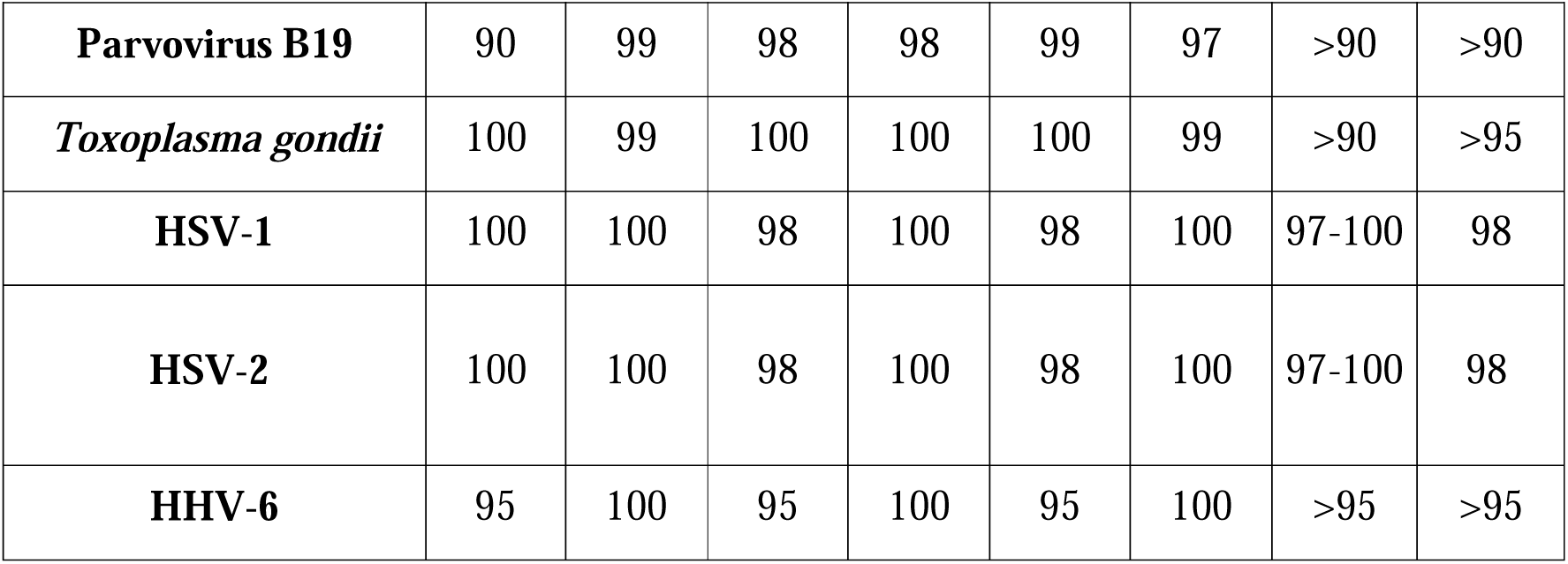
Antigen sensitivities. shows the IgG, IgM, and IgG+IgM sensitivities and specificities obtained using the Vibrant microarray and a comparison with the sensitivities and specificities of the current gold standard for respective pathogens.

## Enhanced IgM Assay

IgM antibodies are the primary antibodies produced by the immune system during infections, but they make up only 5% to 10% of all the circulating antibodies [33]. An in-house IgM assay was developed to remove most IgG antibodies and other non-specific proteins from the serum prior to the IgM immunoassay. This helped to increase the sensitivity and specificity of the assay. Human IgG was removed by incubating the serum with a purified goat anti-human (GAH) IgG Fc fragment and proprietary assay reagents.

## Analytical Performance

The analytical performance of the immunochip was evaluated for precision (repeatability/reproducibility), analytical sensitivity, reportable range, linearity, and matrix equivalency studies. Samples for negatives, low or moderate positives, and high positives were run with duplication to determine the analytical performance metrics. The precision study used a panel of 11 samples and was run over a period of 20 days with 2 duplicates per run and 4 runs per day. The results are tabulated as shown in Supplementary Table 1. Lot to Lot reproducibility was also tested to check for variation in the manufacturing of the pillar plates by running a panel of 11 samples with 5 replicates per run, 3 runs per day over a period of 5 days using 3 manufactured lots. The results are tabulated as shown in Supplementary Table 2. Testing of protein-free serum matrix samples and low antibody concentration samples with 2 replicates per run, 2 runs per day over a period of three days was used to determine analytical sensitivity. The limit of blank (LoB) and limit of quantitation (LoQ) was calculated using the mean and standard deviation of the blank and the low antibody concentration samples as shown in Supplementary Table 3. The linearity and reportable range were verified by running samples with varying levels of antibodies and checking assay recovery, the results are tabulated in Supplementary Table 4. Matrix equivalence studies are shown in Supplementary Table 5. The potential interference of specific endogenous and exogenous substances with the immunochip was evaluated by performing an interfering substance study. The interfering substances tested were 60 mg/dl bilirubin, 100 mg/ml cholesterol, 1000 mg/ml triglycerides, 1000 mg/ml hemoglobin, and 6 g/dl albumin. There was no interference between the immunochip and the substances tested at the mentioned levels.

## Clinical Sensitivity and Specificity

Table 3 provides an overview of the IgG and IgM sensitivities measured by the Vibrant microarray. This is compared with the sensitivities and specificities of the current gold standard tests for the particular pathogen. The Vibrant microarray was able to achieve high sensitivities and specificities when compared with the gold standards for each pathogen. Supplementary Table 7 provides more details on the gold standard diagnostic tests for the pathogens along with the modes of transmission and their endemic regions.

## Evaluating the antigens of *Borrelia burgdorferi*

In this study, individual antigens of *B. burgdorferi* were tested for reactivity with IgG and IgM antibodies (Table 4). The heat map (Figure 3) shows the performance metrics of the different antigens. Testing for Lyme disease since 1994 has been based on conventional two-tiered testing (CTTT) where an enzyme immunoassay (EIA) is followed by a specific immunoblot for a definitive diagnosis. Recently, CTTT has been replaced with a modified two-tier testing (MTTT) in which an EIA using whole cell sonicate is followed by an EIA using C6 peptide. This shows the increasing shift away from blot-based testing to conventional ELISA. MTTT removes the burden of immunoblots which are tedious to run, more expensive and could have subjective interpretation of bands [49]. Complete replacement of immunoblots can be done using a microarray platform such as the one described here. The full data set would be available to the physicians to make a nuanced diagnosis instead of a narrow subset of antigens run on ELISAs.

**Figure 3:**
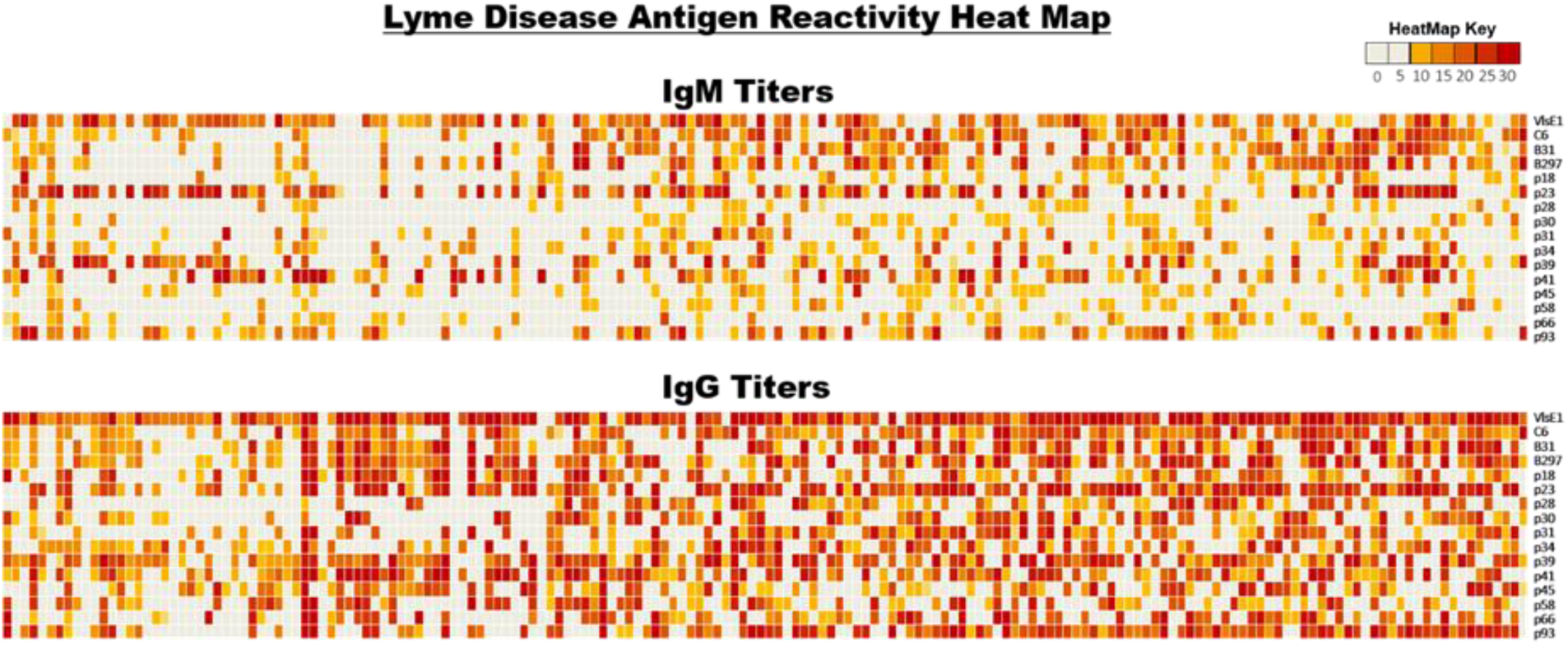
H**e**at **map showing Lyme disease antigen reactivity**. The positivity cutoff for each antigen was set at greater than 10 chemiluminescent units (CU) (shown as yellow or red). The color key is as follows: Red – High positive (CU>20); Yellow, orange – Moderate positive (CU = 10.1-20); White – Negative (CU≤10).

**Table 4.** Individual antigen sensitivities (*B. burgdorferi*) shows the IgG and IgM values for the individual antigens of *Borrelia burgdorferi*

| Antigen | IgM | IgG |
| --- | --- | --- |
| ***VlsE1*** | 61% | 91% |
| ***C6*** | 39% | 60% |
| ***B31*** | 37% | 48% |
| ***B297*** | 36% | 57% |
| ***p18*** | 6% | 41% |
| ***p23*** | 47% | 68% |
| ***p28*** | 4% | 28% |
| ***p30*** | 5% | 35% |
| ***p31*** | 17% | 38% |
| ***p34*** | 16% | 41% |
| ***p39*** | 29% | 66% |
| ***p41*** | 34% | 53% |
| ***p45*** | 5% | 35% |
| ***p58*** | 5% | 35% |
| ***p66*** | 7% | 39% |
| ***p93*** | 32% | 60% |

## Discussion

Ticks are among the most important sources of vector-borne infections in the US [50]. The spread of ticks across the US has been steadily increasing over the past decades. In parallel, the discovery of novel pathogens that are spread by ticks has also seen dramatic increases [51]. Currently, there are 11 major tickborne diseases according to the CDC namely, Lyme disease, babesiosis, ehrlichiosis, Rocky Mountain spotted fever, Southern tick-associated rash illness, tick-borne relapsing fever, tularemia, anaplasmosis, Colorado tick fever, and Powassan encephalitis [52] (https://www.cdc.gov/ticks/diseases/index.html). Patients are rarely tested for all possible infections that could be transmitted via a tick bite [53]. The current diagnostic tests are severely limited in distinguishing various tick-borne infections and several studies have revealed that non-Lyme tick-borne infections are heavily underdiagnosed [54].

Among varied testing options PCR and serology-based assays are reliable and most widely used. PCR has several advantages as it detects pathogenic DNA/RNA which conclusively proves the organism’s presence. It has high specificity and has a high throughput with assay run times of about 2 hours. It can also detect the infection during its early stages [55]. There are however certain drawbacks to testing using PCR, especially with tickborne infections. Pathogens transmitted by ticks may be transient in the blood resulting in false negative PCR results in tick-borne diseases, namely, *B. burgdorferi, R. typhi, T. gondii*, HSV-1, EBV, TBEV, and WNV [23]. PCR testing requires specialised laboratories and equipment for testing. PCR may not detect all strains and variants and is limited to detecting known pathogens [56]. Multiplexing using PCR is limited due to fixed number of analytes that can be parallelly read using PCR instrumentation.

Serology-based testing has several advantages when it comes to tick-borne infection testing. It has the ability to comprehensively assess immune responses and simultaneously detect exposure to multiple pathogens including previous and unresolved infections. Testing two times with a time interval in between can also help diagnose active infections based on altered serum antibody profiles. Simultaneous detection of antibodies against multiple tick-borne pathogens using a single sample and providing a comprehensive view of the patient’s immune response is a key advantage of serology-based multiplex testing [23]. The testing can also be done in resource poor settings with collection using a dried blood spot [22]. Serological testing can diagnose tick-borne diseases even in the later stages when pathogen detection through molecular methods becomes more challenging [23,57]. It also reduces the risk of false negatives [57]. Serological multiplex testing being cost-effective can also contribute to surveillance and epidemiological studies by providing valuable data on the prevalence and distribution of tick-borne diseases, enhancing our understanding of disease dynamics [58]. However, serological studies have their own limitations. Serological testing may not be able to detect early/recent infections. It relies heavily on the timing of sample collection and the host’s immune responses. In certain cases, molecular testing may be needed to confirm serological testing [23]. Despite all this, the benefits of serology testing outweigh its limitations which is why it is recommended by the CDC as the standard of testing for Lyme disease.

Apart from PCR and ELISA serology tests, IFA and culture methods have also been suggested for diagnosing tick-borne infections. Testing using IFA is limited due to a lack of standardized antigenic targets, the subjective establishment of positive thresholds, and cross reactivity. These factors can result in varying accuracy of IFA results across laboratories [23]. Furthermore, Bacterial or viral cultures are not recommended for the diagnosis of tick-borne infections. This is due to the time-consuming nature of the test, the need for special media, and procedures that are only performed at specific laboratories [61].

This study employed a serology-based microarray developed at Vibrant to multiplex Lyme and other tick-borne infections along with a few other infections of interest. The uniqueness of the microarray lies in the application of the immunodominant antigens that eliminate nonspecific binding with high sensitivity needed for accurate diagnosis. Antigens could be evaluated in a multiplex setting to gauge their performance with clinical samples to pick the ideal set of antigens for any infection. To the best of our knowledge, this is the first report on the broad panel of antigens for Lyme disease and its co-infection testing in such a flexible format. The structure of the Vibrant pillar plate is designed to encompass 400 probe chips at each pillar facilitating the detection of an array of co-infections in a single run, saving cost, labor, and time. Further compaction of the chip allows improved performance by enhancing multiplexing and widening its clinical applications. The microarray platform is advantageous over other existing gold standards for tick-borne diseases and was able to overcome several of their limitations. Average time for multitier testing for several tickborne pathogens could take several months whereas the microarray technology takes only about a day to perform [21]. The microarray detected 17 tick-borne and other infections along with Lyme disease with sensitivities and specificities listed in Table 3.

In conclusion, the protein microarray with a multiplex of antigens was validated for Lyme and its co-infections. The impact of simultaneous testing of co-infections leads to focused and efficacious therapeutic recommendations. This approach caters to the diagnostic needs of patients owing to its high sensitivity and specificity, affordable cost, quick availability of results, and low sample volume requirement. Measures for syndromic surveillance, diagnostic preparedness in disease outbreak investigations, personal protection, and education of clinical health professionals and patients could pave the way for controlling tick-borne infections better. As the known repertoire of antigens increases, this flexible microarray format can be customised to include these new antigens. Future editions could also include other infections/agents namely Colorado tick fever, heartland virus, rickettsiosis, Rocky Mountain spotted fever, Southern tick-associated rash illness, tick-borne relapsing fever, and tularemia which can be tested in parallel. Novel antigens for pathogens which may include whole cell sonicates, recombinant proteins or peptide epitopes can be added as the science progresses leading to continuous improvement in diagnostic technology for detecting tick-borne infections.

## Declarations

### Author Contributions

Conception and study design: HKK, JJR, VJ. Performing experiments: KK, TW. Analysis and interpretation: KB, KK. Writing-original draft: HKK, CS, SM. Review and editing: HKK, CS, AJR, RWF, AP, LSB, AC, AF, DR, GKN, LJA, DWA, MAA, DL, CF, PD, MF, MSV, KH, DC, AP, EM, and AN. Sample resources: RWF, AP, LSB, AC, AF, DR, GKN, LJA, DWA, MAA, DL, CF, PD, MF, MSV, KH, DC, AP, EM, and AN. All authors reviewed and approved the final manuscript.

## Data Availability

The data used to support the findings of this study can be acquired from Vibrant America LLC

## Acknowledgment

Vibrant Sciences LLC developed the microarray technology showcased in the publication. All IP associated with the microarray manufacture and diagnostics belongs to Vibrant Sciences. The specific roles of authors are stated in the author contribution section.

## Availability of Data

The data used to support the findings of this study can be acquired from Vibrant America LLC.

